# Preimmunization correlates of protection shared across malaria vaccine trials in adults

**DOI:** 10.1101/2021.08.13.21262031

**Authors:** Maxwell L. Neal, Fergal J. Duffy, Ying Du, John D. Aitchison, Kenneth D. Stuart

## Abstract

Identifying preimmunization biological characteristics that promote an effective vaccine response offers opportunities for illuminating the critical immunological mechanisms that confer vaccine-induced protection, for developing adjuvant strategies, and for tailoring vaccination regimens to individuals or groups. In the context of malaria vaccine research, studying preimmunization correlates of protection can help address the need for a widely-effective malaria vaccine, which remains elusive. In this study, common preimmunization correlates of protection were identified using transcriptomic data from four independent, heterogeneous malaria vaccine trials in adults. Systems-based analyses showed that a moderately elevated inflammatory state prior to immunization was associated with protection against malaria challenge. Functional profiling of protection-associated genes revealed the importance of several inflammatory pathways, including TLR signaling. These findings, which echo previous studies that associated enhanced preimmunization inflammation with protection, illuminate common baseline characteristics that set the stage for an effective vaccine response across diverse malaria vaccine strategies in adults.

## Introduction

Whether a vaccine generates an immune response that results in protection from infection or disease depends in part on the preimmunization status and capacities of the vaccinated individual’s immune system^1^. Identifying preimmunization correlates of protection, therefore, offers opportunities for illuminating the immunological pathways that promote or attenuate protective vaccine responses, for identifying avenues whereby the immune system can be influenced to respond effectively, and for developing personalized approaches that predict vaccine efficacy on an individual or group basis. These opportunities can potentially benefit malaria vaccine development efforts: Although substantial progress has been made developing anti-malaria vaccines, a vaccine that is widely-effective has remained elusive^2–5^, and it remains unclear why malaria vaccines induce a protective response in some individuals but not in others. Exploring the preimmunization correlates of protection in malaria vaccine trials presents an opportunity to identify the immunological conditions that set the stage for vaccine-induced responses that are protective against future infection and to help guide the rational design of more effective vaccines. While the list of studies in this research area is not extensive, some preimmunization correlates of protection have been identified for vaccines against influenza^6–9^, hepatitis B^10–12^, yellow fever^13^ and malaria^14–21^.

To identify common preimmunization correlates of protection across various malaria vaccine trials, we performed transcriptomic analysis of four independent trials: the Immunization via <u>M</u>osquito Bite with <u>R</u>adiation-<u>A</u>ttenuated *P. falciparum* <u>S</u>porozoites (IMRAS) trial^22^, the <u>B</u>agamoyo <u>SP</u>oro<u>Z</u>oite <u>V</u>accination 1 (BSPZV1) trial^23^, the MAL68 RTS,S vaccine trial^24^, and a Chloroquine Prophylaxis and Sporozoites (CPS) trial^25,26^. These trials varied in the vaccines administered (live, radiation-attenuated whole sporozoites, RTS,S subunit vaccine or live sporozoites where establishment of blood stage infection was prevented by chloroquine chemoprophylaxis), the route of administration (intravascular injection, intramuscular injection, or mosquito bite) the country of trial subject recruitment (USA, Tanzania, or The Netherlands), whether subjects were malaria-naïve prior to immunization (USA, The Netherlands) or experienced (Tanzania), and the source of transcriptomic material (whole blood or peripheral blood mononuclear cells - PBMCs). Each trial included a challenge phase, wherein subjects were infected with infectious *P. falciparum* to determine whether the subject mounted a vaccine response protective against malaria. To our knowledge, this study is the first to investigate preimmunization correlates of protection from transcriptomics across multiple, heterogenous malaria vaccine trials in adults. Despite the wide-ranging clinical trial conditions, our results indicate that protection against post-immunization malaria challenge is associated with innate immune system activation and a moderately elevated inflammatory state prior to immunization, including higher expression in genes associated with TLR signaling and other inflammatory pathways. These results echo molecular- and pathway-level preimmunization correlates of malaria protection derived from previous analyses on narrower sets of clinical trials^19,21^. They highlight specific immunological pathways that may play critical roles in the development of effective malaria vaccine-elicited responses and offer potential targets for manipulating the immune system into a state that promotes such responses.

## Results

### Correlates of protection among pooled preimmunization transcriptomes

Preimmunization transcriptomes were pooled from the four malaria vaccine trials and consisted of 84 samples (Table 1, Methods). A differential expression analysis comparing the transcriptomes of all protected participants (*n*=38) and all non-protected participants (*n*=46) pooled from the trials revealed no genes with statistically-significant differences in transcript abundance using an FDR-adjusted *P*-value cutoff of 0.1. To test for differences among functionally-related *groups* of genes, we performed Gene Set Enrichment Analysis (GSEA)^27^ to identify statistically-significant overrepresentation of predefined gene sets including immunological blood transcriptional modules^28,29^ and the MSigDB Hallmark gene sets^30^. GSEA results on genes ranked according to their DESeq2 Wald test statistic revealed significant differences in transcript abundances (FDR-adjusted *P*-value < 0.05) among 207 gene sets. Gene sets enriched for higher abundance transcripts in protected subjects were predominantly associated with inflammation and inflammatory signaling pathways, myeloid lineage cells including monocytes and neutrophils, coagulation, antigen presentation, cell death and apoptosis, the endoplasmic reticulum, the extracellular matrix, and the complement system (Fig. 1). Gene sets enriched for lower abundance transcripts in protected subjects were predominantly associated with the cell cycle, protein synthesis, interferon, T cells, mitochondria stress/respiration, and NK cells. As illustrated in Fig. 2, leading-edge genes (those primarily responsible for the significant enrichment scores due to non-random grouping at the tails of a ranked list) from gene sets with the highest and lowest normalized enrichment scores (NESs) showed moderate differences in transcript abundance between protected and non-protected subjects. For example, the top 10 leading-edge genes in Fig. 2a showed a mean log2 fold-change of 0.41 ± 0.22 (SD) between protected and non-protected subjects. Our analysis revealed that, when comparing protected and non-protected subjects, differences in transcript abundance of gene *sets* showed statistical significance, whereas differences at the individual gene level did not.

**Table 1.** Characteristics of the four clinical trials that generated transcriptomic data used to identify preimmunization correlates of protection.

| Trial name<br>(ClinicalTrials.gov ID) | Immunogen | Recruitment<br>country | Sample<br>source | N<br>preimmunization<br>samples | Prior<br>malaria<br>exposure | N female /<br>male | N challenge-protected /<br>non-protected |
| --- | --- | --- | --- | --- | --- | --- | --- |
| IMRAS<br>*Immunization via Mosquito Bite with*<br>*Radiation-Attenuated P. falciparum*<br>*Sporozoites*<br>(NCT01994525) | Radiation-attenuated<br>sporozoites via mosquito bite | U.S.A. | Whole blood | 11 | Naïve | 1 / 10 | 6 / 5 |
| BSPZV1<br>*Bagamoyo SPoroZoitE Vaccination 1*<br>(NCT02132299) | Radiation-attenuated<br>sporozoites via IV injection | Tanzania | Whole blood | 22 | Non-naïve | 0 / 22 | 8 / 14 |
| MAL68<br>(NCT01366534) | RTS,S/A01 ± Ad35.CS.01<br>vaccine via injection | U.S.A. | PBMCs | 45 | Naïve | 20 / 25 | 21 / 24 |
| CPS<br>*Chloroquine Prophylaxis and Sporozoites*<br>(NCT01218893) | Sporozoites via mosquito bite<br>(under chloroquine cover) | The<br>Netherlands | Whole blood | 7 | Naïve | 3 / 4 | 3 / 4 |

**Figure 1.**
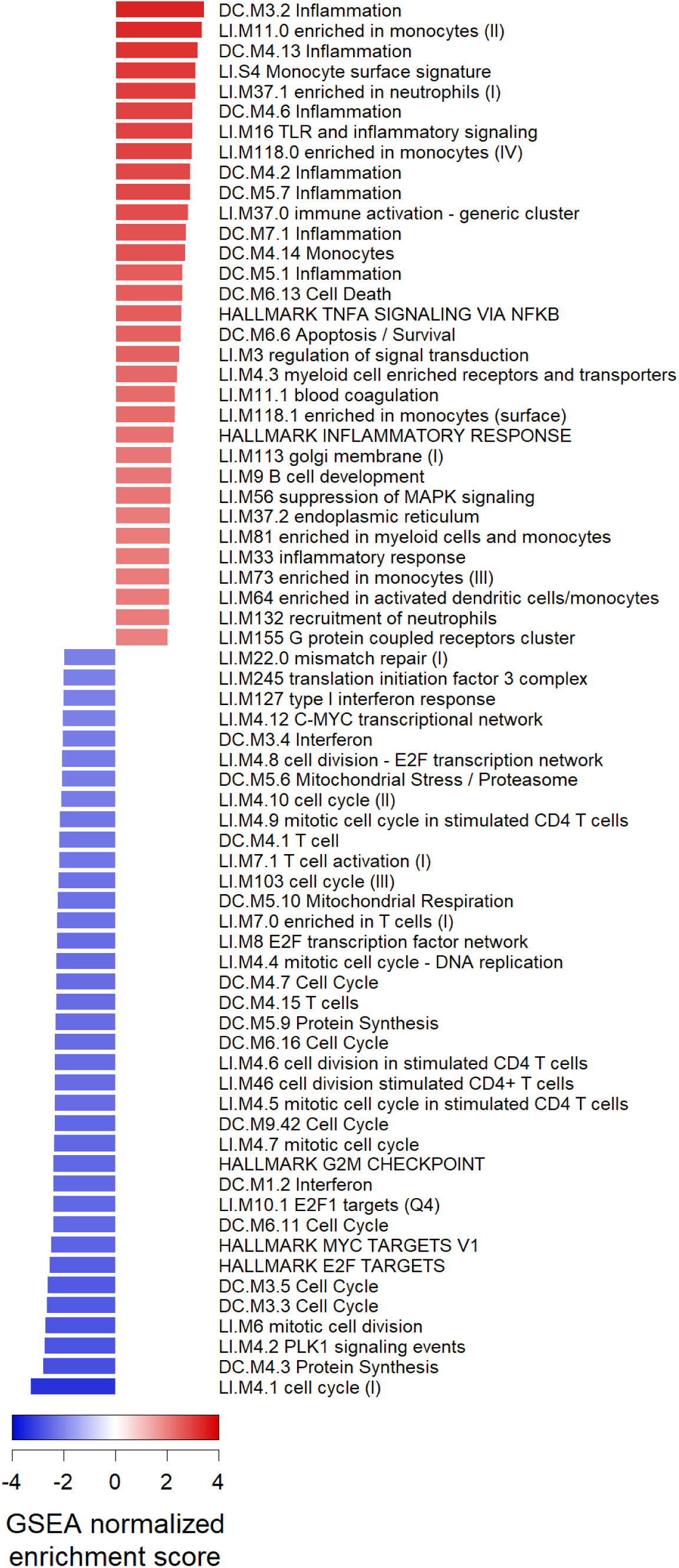
GSEA results on the pooled set of preimmunization transcriptomes from four malaria vaccine trials. Gene sets with positive GSEA normalized enrichment scores indicate they are significantly enriched for genes with higher transcript abundance in protected subjects; negative scores indicate significant enrichment for genes with lower transcript abundance in protected subjects. Gene sets shown have absolute scores > 2.0 and FDR-adjusted *P*-values < 0.05. Sets named “Undetermined” or “TBA” are not shown.

**Figure 2.**
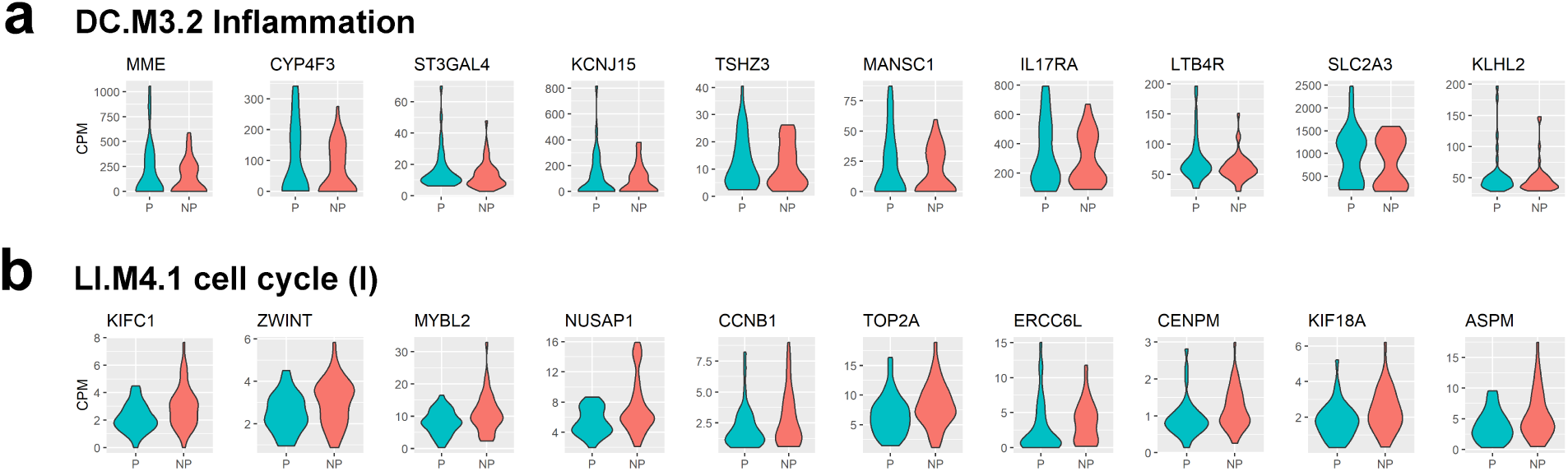
Violin plots comparing transcript abundance of GSEA-derived leading-edge genes in protected (P) and non-protected (NP) subjects pooled from four malaria vaccine trials. **a** Transcript abundance in counts per million (CPM) for the top ten leading-edge genes in *DC*.*M3*.*2 Inflammation*, the gene set showing the highest significant normalized enrichment score in the GSEA analysis (indicating significantly higher abundances in protected subjects). **b** Transcript abundance for the top ten leading-edge genes in *LI*.*M4*.*1 cell cycle (I)*, the gene set showing the lowest significant normalized enrichment score in the analysis (indicating significantly lower abundances in protected subjects).

### Correlates of protection shared across individual trials

The analyses on the pooled set of preimmunization transcriptomes includes all samples aggregated across all four trials. To perform a more conservative analysis that limits correlates driven primarily by individual trials, we computed differential expression and performed GSEA for each trial separately, then identified genes and gene sets that consistently showed significant differences between protected and non-protected subjects across trials. As with the differential expression analysis on the pooled transcriptomes, the number of transcripts showing significant differences in abundance between protected and non-protected subjects was low in each trial (*N* = 2 for IMRAS, *N* = 0 for BSPZV1 and MAL68, *N* = 14 for CPS; FDR-adjusted *P*-value < 0.1). To identify differences at the level of gene sets, we then performed the same type of GSEA as performed with the pooled samples. From the GSEA results on each trial, we then identified gene sets that showed significant positive enrichment scores consistently across all trials or consistently showed significant negative scores. Fourteen gene sets showed directionally-consistent, significant enrichment for genes with higher expression in protected subjects across trials, and two sets showed directionally-consistent, significant enrichment for genes with lower expression in protected subjects across trials (Fig. 3).

**Figure 3.**
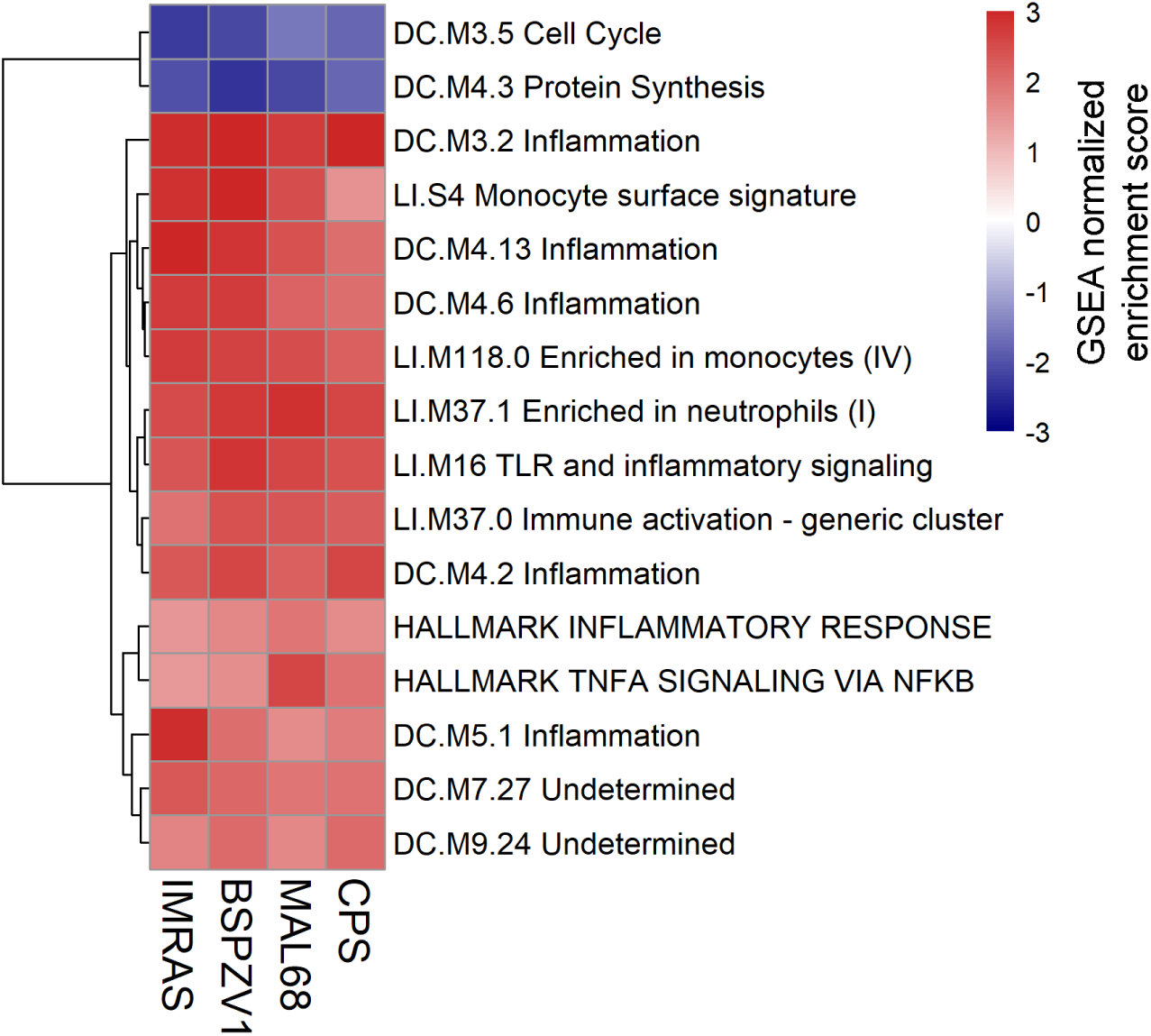
**Heatmap of GSEA normalized enrichment scores for gene sets showing directionally-consistent, significant enrichment for gene transcripts with higher abundance in protected subjects across malaria vaccine trials or lower abundance across trials**.

The 14 gene sets showing consistently higher transcript levels in protected subjects prior to immunization were predominantly associated with inflammatory responses, monocytes, and neutrophils. The two gene sets showing consistently lower transcript levels in protected subjects were associated with the cell cycle and protein synthesis. To determine the probability of seeing 16 gene sets significantly enriched across all four trials with consistent directionality in their enrichment scores by chance, we randomly assigned protected/non-protected status to each trial subject, re-computed differential expression between protected and non-protected subjects, performed GSEA on the ranked DESeq2 results, and then recorded the number of gene sets showing directionally-consistent, significant enrichment in all four trials. This was done in a manner that preserved the original number of protected and non-protected subjects in each trial. We performed 10,000 iterations of this procedure, each of which used a unique set of protection assignments and found that the probability of seeing 16 or more directionally-consistent, significantly-enriched gene sets appear in all four trials was less than 0.007.

### Pathway analysis of leading-edge genes common across trials

While the gene sets used in our GSEA analysis are useful for characterizing gene groups according to their more general biological functions, they do not represent specific mechanistic molecular pathways. To identify such pathways that were consistently associated with protection at the preimmunization timepoint, we performed an Ingenuity Pathway Analysis (IPA)^31^ on the set of common leading-edge genes collected from the gene sets in Fig. 3 that showed directionally-consistent, significant enrichment across vaccine trials. For each of these 16 functional gene sets, we collected the genes that appeared as GSEA leading-edge genes for that set in all four trials. Across these 16 gene sets, we found 98 such genes (Supplementary Table S1). Functionally profiling these genes collectively with IPA yielded significant enrichment (Fisher’s exact test, FDR-adjusted *P*-value < 0.05) for 28 IPA Canonical Pathways (Fig. 4). Reflecting the GSEA results, the 28 pathways were primarily associated with inflammation, including many pathways in which Toll-like receptors (TLRs) and MYD88 participate. For each of the 98 genes input to IPA for this analysis, we computed the mean log_2_ fold-change differences in transcript abundance between protected and non-protected subjects across trials, and these values were used by IPA to identify which of the 28 Canonical Pathways showed a consistent activation or inactivation profile. The Canonical Pathway with the highest enrichment score that also showed a consistent activation or inactivation profile was the Toll-like Receptor Signaling pathway. Fig. 5 shows the IPA diagram for this pathway, highlighting the overlap between members of the 98-gene list and the pathway’s molecular participants as well as the mean abundance differences of those overlapping participants when comparing protected to non-protected trial subjects.

**Figure 4.**
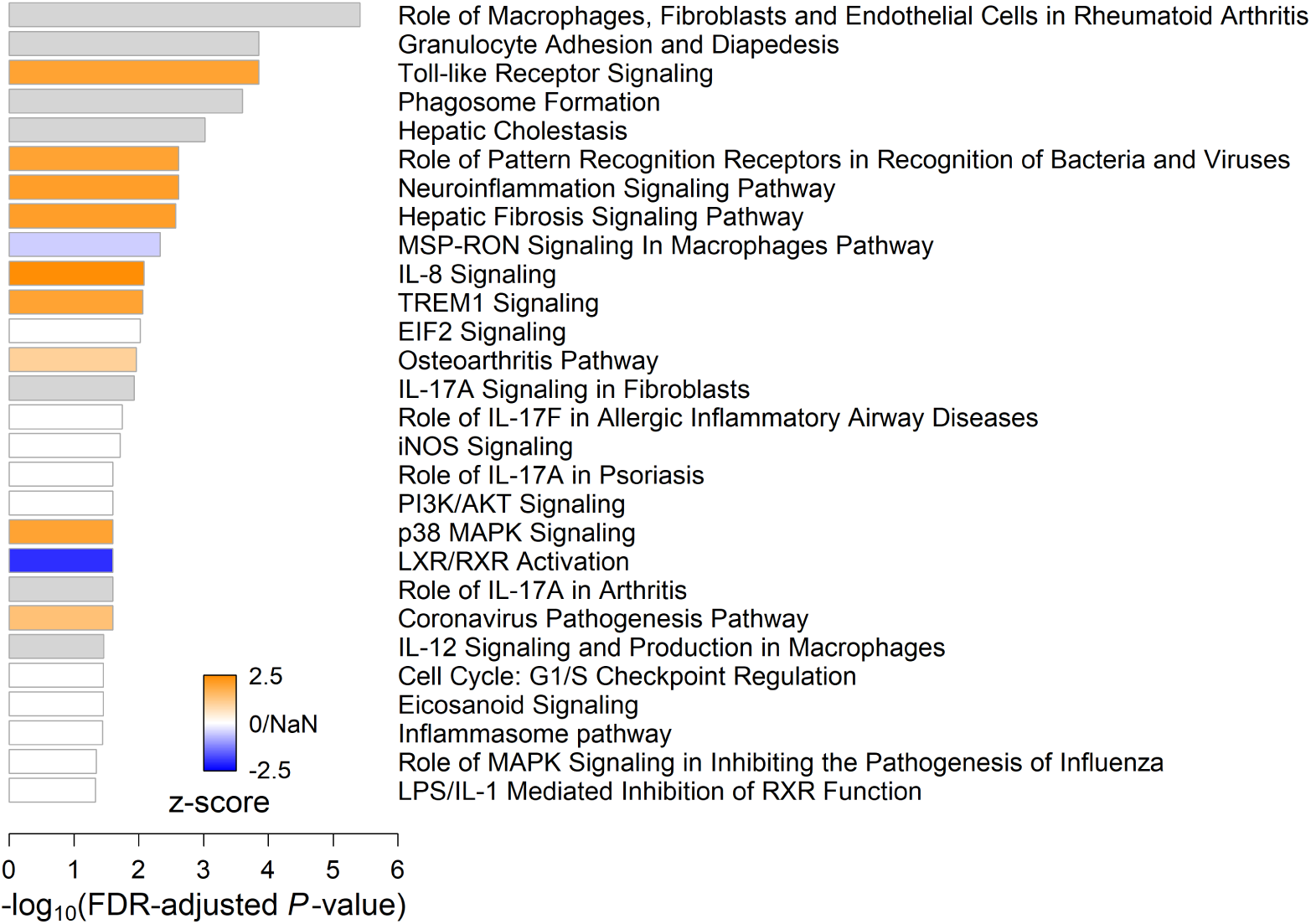
**IPA Canonical Pathways analysis on common leading-edge genes from gene sets that showed directionally-consistent, significant GSEA enrichment scores across trials**. Bar width indicates the strength of an IPA Canonical Pathway’s enrichment for the leading-edge genes. Bar color indicates the IPA z-score, which is the degree to which the pathway is activated (orange) or inactivated (blue) based on mean log_2_ fold-changes in transcript abundance between protected and non-protected subjects for leading-edge genes found in the pathway. White bars indicate an ambiguous activation state (z = 0) or pathways ineligible for activation analysis because the overlap between common leading-edge genes and pathway participants used to compute the z-score was less than four (z-score = NaN). Gray bars indicate that there is insufficient evidence in the IPA Knowledge Base to confidently predict an activation state for the pathway.

**Figure 5.**
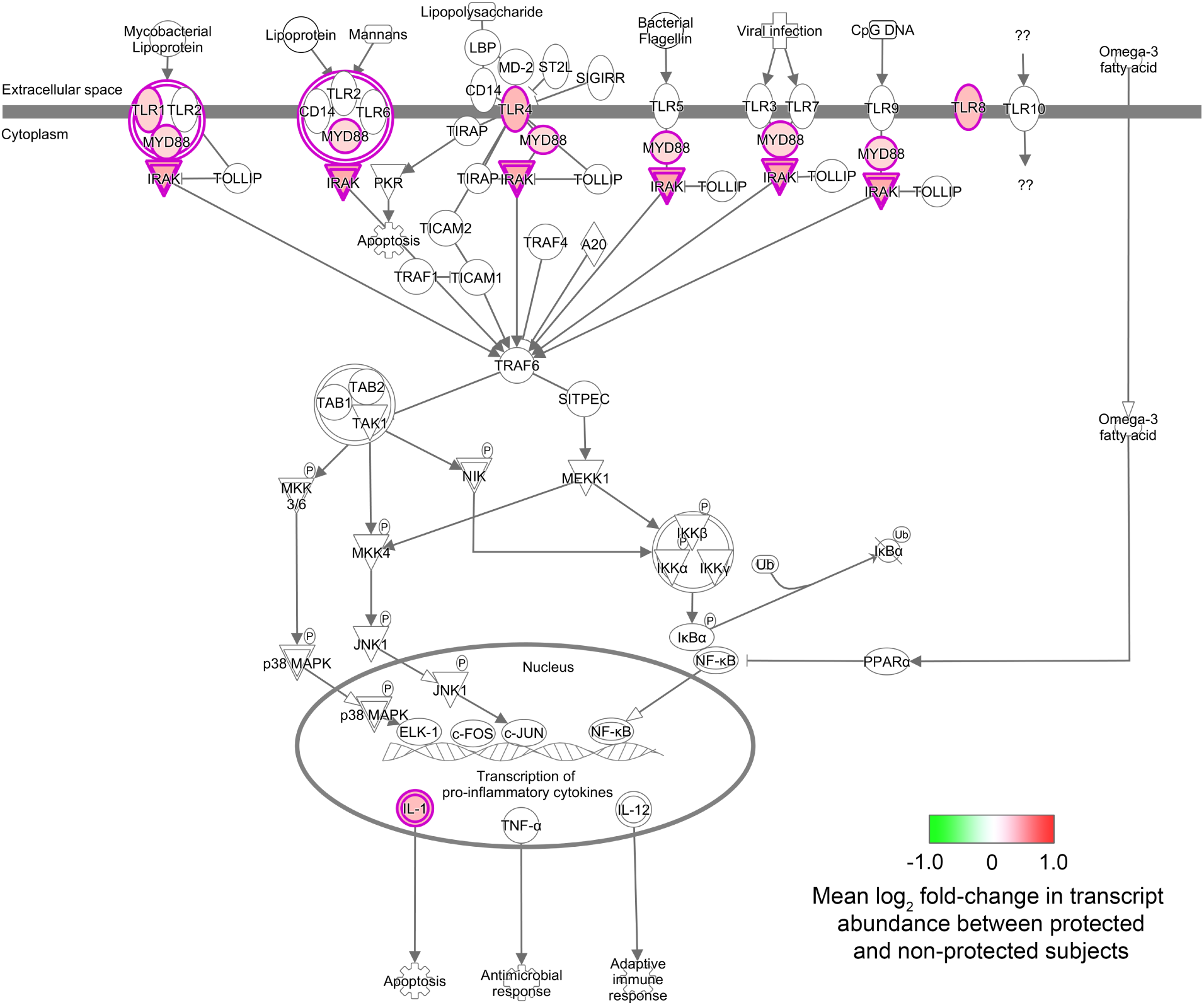
IPA network diagram of the Toll-like Receptor Signaling pathway, illustrating higher preimmunization activity in protected subjects. The pathway shows enrichment for six protection-associated leading-edge gene transcripts found across vaccine trials, indicated by icons with red borders. Icon fill color indicates the log2 fold-changes in transcript abundance when comparing protected to non-protected subjects, averaged across the four vaccine trials.

In addition to the Canonical Pathways analysis, we also performed an IPA Upstream Analysis, which provides hypotheses about which molecular perturbations might cause the expression differences observed in the 98-gene list. In the context of this study, this analysis can help identify pathway activators and inactivators (naturally-occurring or synthetic) with the potential to shift an individual’s immunological state in the direction that is more consistent with a protective response to immunization. The molecules with high IPA activation z-scores (≥ 2.0) showing significant FDR-adjusted *P*-values in this analysis included cytokines with proinflammatory activity (type I and type II interferons, IL1B, TNF, IL17A, IL33, IL18, IL6, IL12, IL1A, IL5), cytokine groups (IL12, IL1), lipopolysaccharides, colony stimulating factors (CSF2 and the pharmaceutical analog of CSF3 filgrastim), transcription regulators (the NFκB complex, CEBPA, SPI1, TCF7L2, STAT3, RELA, IRF7), Toll-like receptors (TLR2, TLR4) as well as adapter proteins that participate in their signaling pathways (TICAM1, MYD88), other immunomodulators (tretinoin, ethanol, SPI1, TGM2, poly rI:rC-RNA, mycophenolic acid, POU2F2, KITLG), the protein synthesis regulator LARP1, thrombin, APP, signaling pathway enzymes and ligands (PARP1, PI3K, KITLG, the p38 MAPK group), the PDGF-BB complex, the growth hormone protein group, the C11orf95-RELA gene fusion product, and ZBTB10. Based on the IPA knowledge base, increased activity of these molecules would shift expression of the 98 leading-edge genes in a manner consistent with their mean expression differences between protected and non-protected subjects across trials. The upstream molecules with low IPA activation z-scores (≤ -2.0) showing significance included the p38 MAPK inhibitor SB203580, the progesterone antagonist mifepristone, miR-155-5p, the alpha catenin protein group, the transcription factor MLXIPL, the transcription inhibitor actinomycin D, DIO3 and DUSP1, the gene fusion product ETV6-RUNX1, the immunosuppressant cyclosporin A, and the corticosteroid budesonide. Decreased activity of these molecules is predicted to induce expression changes in the 98 leading-edge genes consistent with those observed between protected and non-protected subjects. The full set of Upstream Analysis results are provided in Supplementary Table S2.

### Discriminatory power of protection-associated genes and gene sets

The protection-associated genes and gene sets common across trials suggest there are transcriptomic features that might be used in classifiers that discriminate, on an individual basis, which vaccinees will mount a protective immune response to immunization and which will not. To assess the discriminatory power of these features, we developed and tested various scores based on transcript abundances (counts per million) of the 98 protection-associated, leading-edge genes described above as well as mean transcript levels corresponding to those genes within protection-associated gene sets. We also tested the discriminatory power of scores based on transcript abundance *ratios* between protection-associated genes and between gene sets. For all scoring strategies, individual scores were computed for each trial participant based on their preimmunization transcriptomic profile. We then assessed the discriminatory power of the score by varying the threshold used to classify individuals as protected or non-protected and then generating a receiver operating characteristic (ROC) curve from the sensitivity and specificity of the classifiers. We quantified the discriminatory power of each score based on the area under the curve (AUC) of the ROC results. We found that the ratio of mean leading-edge gene expression in the *HALLMARK TNFA SIGNALING VIA NFKB* gene set to that of the *DC*.*M4*.*3 Protein synthesis* gene set produced a score with the highest discriminatory power (AUC = 0.73, 95% confidence interval 0.61-0.84, Fig. 6a). A Mann–Whitney U test showed that this score was significantly different between the protected and non-protected subjects pooled from all four trials (FDR-adjusted *P*-value = 0.034, Fig. 6b). The score incorporates expression information from eight members of the *HALLMARK TNFA SIGNALING VIA NFKB* gene set (BCL3, BTG1, CCRL2, CEBPD, CXCL1, PFKFB3, PTGS2, SLC2A3) and seven from the *DC*.*M4*.*3 Protein synthesis* set (RPL4, RPL5, RPL6, RPS3, RPS14, RPS18, TOMM7).

**Figure 6.**
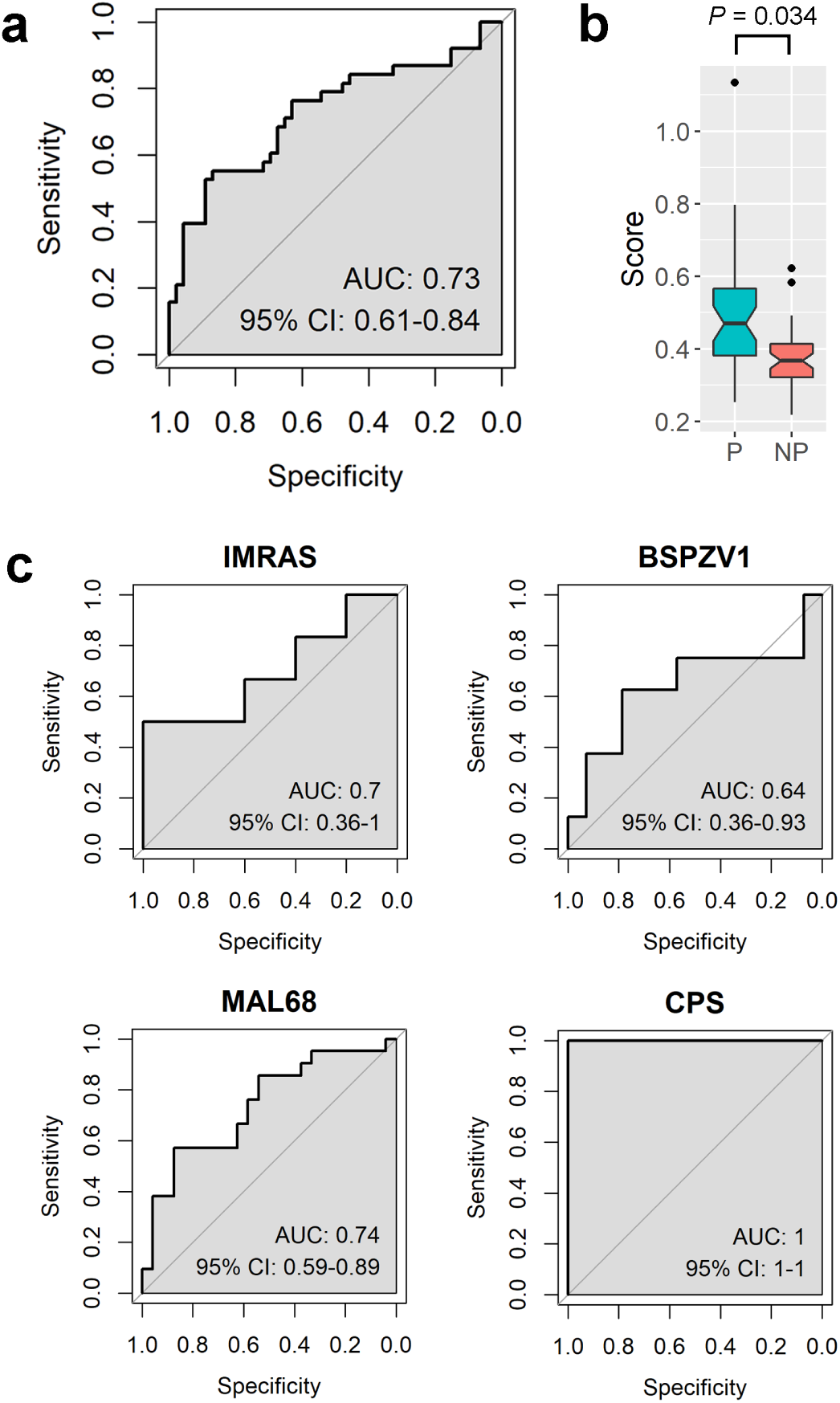
Discriminatory power of the best-performing preimmunization score for classifying protected and non-protected trial subjects. **a** Receiver operating characteristic (ROC) curve showing performance of the score across 84 subjects pooled across trials. **b** Boxplots showing score distributions among subjects who were protected (P) following malaria challenge and those who were not (NP). **c** ROC curves showing performance of the score within each individual trial. (AUC: area under the curve; CI: confidence interval).

To test whether the discriminatory power of the best-performing score might be biased by specific trials in our study, we computed separate ROC curves for each trial using only their participating subjects and their corresponding scores. We found that the score showed comparable discriminatory power across trials (Fig. 6c). The score’s performance was poorest in the BSPZV1 cohort (AUC=0.64, 95% confidence interval 0.36-0.93) and highest in the CPS cohort where it perfectly discriminated between protected and non-protected subjects.

## Discussion

Our findings indicate that a moderately elevated preimmunization inflammatory state, as revealed by RNA-seq transcriptomic measurements on whole blood or PBMCs, is associated with a protective response to malaria challenge following a variety of immunization strategies in adults. A heightened preimmunization inflammatory state has previously been linked to vaccine-induced protection against influenza in non-elderly adults^6^, and our results also link this finding to the malaria vaccination strategies analyzed here. Our findings contrast with studies investigating correlates of protection following hepatitis B immunization where innate immune activation prior to immunization has been linked to non-protection in adults^10–12^.

Across the four trials we analyzed, transcripts from several inflammatory and innate response gene sets trended higher in protected subjects, and this trend was significant when treating gene sets as the unit of analysis. Analyses at the level of individual transcripts did not reveal significant differences between protected and non-protected subjects across trials. Thus, the expression differences we observed between these groups link protection to a moderately higher activation of inflammatory and innate response processes. While moderate, these differences are substantial enough to have potential utility in metrics that predict whether an individual subject will mount a protective vaccine response prior to immunization (Fig. 6). In future work, metrics based on the discriminatory transcriptomic features identified here might be combined with other protection-associated preimmunization measurements (e.g., cell type abundances) to create robust, accurate classifiers suitable for identifying individuals at risk for a poor vaccine response in a clinical setting.

The set of common leading-edge genes gathered from our GSEA results showed enrichment for more fine-grained signaling pathways, most of which included TLR4 as a participant. These results echo previous findings by Moncunill, et al.^19^ who identified TLR4 as a member of several gene signatures predictive of protection in RTS,S-vaccinated children. These signatures were derived from expression changes in preimmunization PBMCs that were stimulated with CSP and/or *P. falciparum*-infected erythrocytes. Their results suggested that higher preimmunization TLR4 expression may help the immune system respond more effectively to the RTS,S vaccine. As noted by these authors, the LPS-derived chemical monophosphoryl lipid A (MPL) used as an adjuvant in combination with the subunit RTS,S vaccine is a TLR4 agonist^19,32,33^. Our results, which link higher preimmunization expression of TLR1, TLR4 and TLR8 to protection, also suggest a role for TLR agonists as adjuvants in the development of a protective vaccine response.

Moncunill, et al. also found preimmunization signatures of protection in a separate trial where adults were immunized with fresh sporozoites delivered via mosquito bite under chloroquine prophylaxis. The best performing preimmunization signature derived from CSP-stimulated PBMCs for this trial linked protection with proinflammatory activity of participants in the canonical and non-canonical NFκB pathways. These pathway participants included CSF2 (also known as GM-CSF), and our IPA Upstream Analysis identified CSF2, RELA, and NFκB complex activation as potential upstream perturbations contributing to the increased expression and transcript abundance differences we observed between protected and non-protected subjects. Also included in the CPS signature is ITGA2, an integrin that contributes to p38 MAPK signaling which in turn controls the activity of the RELA NFκB subunit. The p38 MAPK signaling pathway was also enriched for leading-edge genes associated with protection across trials in our IPA Canonical Pathways analysis (Fig. 4). The leading-edge genes participating in this pathway included CREB5, IL18RAP, IL1RN and IRAK3, all of which showed elevated expression in protected subjects across trials. These results suggest that, although we did not see overlap between our set of 98 leading-edge genes and the genes in the preimmunization signature associated with CSP-stimulation in the chloroquine prophylaxis trial analyzed by Moncunill, et al., NFκB-mediated inflammation pathways appear to be a critical determinant of protection in both studies.

An important difference between the Moncunill, et al. study and ours is that our transcriptomic measurements were taken from unstimulated tissues and therefore present snapshots of subjects’ preimmunization immunological *states* whereas the Moncunill, et al. measurements reflect immunological *responses* to antigen stimulation. Taken together, these results associate protection with an innate immune system that is simultaneously more activated prior to immunization and more capable of responding to immunization. These results may be driven by higher-than-average innate immune functionality in protected subjects or lower-than-average functionality in non-protected subjects, or a combination of both. A recent report by Yap, et al.^21^ provides evidence suggesting preimmunization correlates of protection may be driven by actively decreased inflammatory functions in non-protected subjects. The authors report that higher baseline expression of T cell inhibitory ligands CTLA-4 and TIM-3 in CD4^+^ T cells is associated with a “slow-responder” phenotype in adults that is less likely to result in sterile protection following immunization via controlled human malaria infection under chloroquine prophylaxis (CHMI-CPS). Increases in these inhibitory ligands can result from mechanisms that protect against inflammation-induced damage (for example in response to chronic parasitemia in malaria-endemic areas). The authors argue that these inhibitory ligands may interfere with NFκB signaling, which is preferentially activated in subjects who mount a protective response to this type of immunization, and this leads to insufficient production of IFN-γ. Rapid induction of IFN-γ following immunization is a characteristic of a “fast-responder” phenotype in CHMI-CPS trials that is more likely to result in sterile protection. Consistent with these findings, our IPA Upstream Analysis identified increased IFN-γ as a perturbation that would generate expression changes consistent with those observed between protected and non-protected individuals. IFN-γ had the highest z-score in this analysis, indicating strong agreement between a hypothetical increase in IFN-γ and our observed expression patterns showing higher inflammation in protected subjects prior to immunization. Given this consistency between our findings and those of Yap, et al., it may be that non-protected individuals in our study had a less successful immunization response due to protective inflammatory inhibition. However, the group of individuals we analyzed was heterogeneous; therefore, we cannot rule out attributing protection to higher-than-average innate responses among protected subjects. Furthermore, these two interpretations are not mutually exclusive: our results may simultaneously reflect immuno-suppressive factors in non-protected subjects and immuno-activating factors in protected subjects.

The results presented here suggest that assessing malaria vaccinee’s preimmunization status may lead to important correlates of protection and tailoring immunization regimes accordingly may emerge as an important element to ensuring a protective response to immunization. Moreover, our results also offer potential pharmacological interventions that may coax an individual’s immune state into one that is primed to respond effectively to immunization.

## Methods

### Transcriptomic data sources

Transcriptomic data were aggregated from adult participants in four malaria vaccine trials that included RNA-seq measurements at preimmunization timepoints and an assessment of protection against infectious *P. falciparum* following immunization. The trials, described below, are summarized in Table 1. Each trial investigated a different malaria vaccine candidate and, together, represent cohorts from malaria-naïve and non-naïve populations. From these trials, 85 preimmunization transcriptomic profiles were available to investigate correlates of protection.

#### Trial 1: IMRAS

The IMRAS trial^22^ (ClinicalTrials.gov identifier NCT01994525: https://clinicaltrials.gov/ct2/show/NCT01994525) was performed in malaria-naïve participants recruited from the United States. The trial assessed the safety, tolerability, and biomarkers of protection of immunization via radiation-attenuated sporozoites delivered by infectious mosquito bites. Preimmunization transcriptomics data were obtained from whole blood samples collected on the day of the first immunization from 11 immunized participants in the trial’s Cohort 1, each of whom received five doses of radiation-attenuated *P. falciparum* via mosquito bite. Three weeks after the fifth immunization, participants were challenged via *P. falciparum*-infected mosquito bites. To facilitate the identification of correlates of protection, the IMRAS immunization regimen was designed to achieve approximately 50% sterile protection in immunized participants: 6 out of the 11 immunized participants (54%) in Cohort 1 were protected following challenge.

#### Trial 2: BSPZV1

The BSPZV1 trial^23^ (ClinicalTrials.gov identifier NCT02132299: https://clinicaltrials.gov/ct2/show/NCT02132299) was performed in male, malaria-exposed recruits from Tanzania. The trial was designed to determine if protective immune responses against *P. falciparum* infection could be generated in a relatively non-immune population within a malaria-endemic country through immunization with aseptic, cryopreserved, live, radiation-attenuated *P. falciparum* sporozoites delivered intravenously (IV). Preimmunization transcriptomic data from 22 immunized participants were obtained from whole blood samples collected on the day of the first immunization. Each participant received five immunizations and, three weeks after the last, were challenged with infectious, aseptic, cryopreserved *P. falciparum* sporozoites delivered IV.

#### Trial 3: MAL68

The MAL68 trial^24^ (ClinicalTrials.gov identifier NCT01366534: https://clinicaltrials.gov/ct2/show/NCT01366534) compared the ability of two malaria vaccination regimens to elicit protection against malaria challenge in malaria-naïve recruits from the United States. The first regimen consisted of three doses of the RTS,S/AS01 vaccine, which combines a recombinant protein encoding part of the malaria parasite’s circumsporozoite (CSP) protein, the hepatitis B surface antigen, and the AS01 adjuvant. This adjuvant includes the Toll-like receptor 4 (TLR4) agonist 3-*O*-desacyl-4′-monophosphoryl lipid A (MPL) and the QS-21 saponin^32^. The second regimen was the same as the first except a different vaccine, Ad35.CS.01, was used for the first immunization. Ad35.CS.01 consists of replication-deficient adenovirus 35 expressing recombinant CSP. Preimmunization transcriptomics from 45 immunized participants (21 from the first regimen, 24 from the second) were obtained from PBMCs collected on the day of the first immunization. Participants were challenged with *P. falciparum*-infected mosquitos two weeks following the third immunization.

#### Trial 4: CPS

The CPS trial^25,26^ (ClinicalTrials.gov identifier NCT01218893: https://clinicaltrials.gov/ct2/show/NCT01218893) investigated the influence of dosing level on the development of protection against malaria challenge when immunizing malaria-naïve participants using *P. falciparum*-infected mosquito bites obtained under chloroquine prophylaxis. Participants were recruited in The Netherlands and received three immunizations separated by 4-week intervals. They were challenged 19 weeks after the last immunization by exposure to bites from five mosquitoes infected with a homologous *P. falciparum* strain. Preimmunization transcriptomics were obtained from whole blood samples collected one week prior to the first immunization and were from seven members of the study group immunized with 15 mosquito bites total.

### Transcriptomic analyses

Raw RNA-seq sequencing data were aligned to the hg19 human reference genome using a previously described pipeline^34^. Briefly, read pairs were adjusted to set base calls with phred scores less than 5 to “N”, and read pairs for which either end had less than 30 unambiguous base calls were removed. The latter step indirectly removes pairs containing mostly adaptor sequences. Read pairs were then aligned to the genome using STAR^35^ version 2.3.1 and gene counts were computed using HTSeq^36^ version 0.6.0.

Differential expression analyses comparing preimmunization sample counts from challenge-protected trial subjects to non-protected subjects were performed using DESeq2^37^ version 1.28.0. Principal Component Analysis (PCA) was used to identify outliers among transcriptomic samples: If a sample’s first or second principal component value was more than three standard deviations from the corresponding principal component’s mean, they were excluded from downstream analyses. One sample from the CPS trial was identified as an outlier based on PCA. To account for batch effects among trials, we used a DESeq2 design formula that included a trial term as well as a protected/non-protected classification term. Differential expression analyses performed on samples from the same trial only included the latter term.

Gene Set Enrichment Analysis (GSEA)^27^ was performed on gene lists ranked by the DESeq2 Wald test statistic using the fgsea package^38^ version 1.14.0 in R. Gene sets used for GSEA were obtained through the tmod R package^39^ and the MSigDB Hallmark gene set^30^ download site (http://www.gsea-msigdb.org/gsea/msigdb/genesets.jsp?collection=H). Ingenuity Pathway Analysis (IPA) content version 62089861 was used to generate all IPA-based results. Receiver operating characteristic analyses were performed using the pROC package^40^ in R. For all statistical tests, FDR-adjusted *P*-values (Benjamini-Hochberg method^41^) less than 0.05 were considered significant unless otherwise indicated.

## Data Availability

Raw sequencing data for the MAL68 trial is publicly available through Sequence Read Archive BioProject PRJNA401870. Raw sequencing data for the CPS trial is publicly available at Sequence Read Archive BioProject PRJNA381264. Data for the IMRAS and BSPZV1 trials will be available through the ImmPort portal (immport.org).

## Acknowledgements

We would like to thank Nana Minkah for helpful discussions regarding preimmunization correlates, and all volunteers who participated in the clinical trials that were analyzed.

## Author contributions

M.L.N. performed all analyses, interpreted findings, and prepared the manuscript. F.J.D. identified and obtained transcriptomic datasets for analysis, generated counts from sequencing data, helped guide analyses, and edited the manuscript. Y.D. generated counts from sequencing data and edited the manuscript. J.D.A. and K.D.S. conceived the study, interpreted findings, and edited the manuscript.

